# The implementation and outcome of a 2-year prospective audit and feedback based antimicrobial stewardship program at a private tertiary care hospital

**DOI:** 10.1101/2021.01.29.21250434

**Authors:** Pooja Thakkar, Tanu Singhal, Sweta Shah, Rohit Bhavsar, Shweta Ladi, Roshan Elizabeth John, Rubina Chavhan, Reshma Naik

**Affiliations:** Kokilaben Dhirubhai Ambani Hospital and Medical Research Institute

**Keywords:** antimicrobial stewardship, prospective audit and feedback, antimicrobial resistance, antibiotics

## Abstract

**Purpose:** Antimicrobial resistance has emerged as a major public health problem with India being one of the worst affected nations. Hence effective antimicrobial stewardship programs (AMSP) are needed. We report the design, implementation and results of a prospective audit and feedback based AMSP at a private tertiary care hospital.

**Methods:** During the study period – January 2018 to December 2019 – the prescription of restricted antimicrobials required the filling of a justification form which was reviewed by the antimicrobial stewardship committee (AMSC) at 48-72 hours. Patients in whom the restricted antimicrobial was stopped earlier than 48 hours were not applicable for review. The eligible prescriptions were judged as justified/unjustified by AMSC based on the patient’s clinical and previous antimicrobial history, course and results of investigations/ cultures, and communicated to the treating team. Compliance to the recommendations of the AMSC was measured. Days of therapy for each restricted antimicrobial/1000 patient days was calculated. Colistin resistance rates in pathogens causing central line associated blood stream infections were compared with previous years.

**Results:** A total of 2397 restricted antimicrobials in 1366 patients were prescribed in the study period of which 1801 prescriptions were applicable for review (75%). Overall, 1.4% of admitted patients were prescribed restricted antimicrobials. The total days of therapy with restricted antimicrobials was 41.5/1000 patient days. The AMSC committee adjudged 12.5% of prescriptions as unjustified and recommendations for de-escalation were accepted in 89%. There was no significant difference in any of the study outcomes between 2018 and 2019. Colistin resistance rates in CLABSI remained stable as compared to previous years.

**Conclusion:** The prospective audit and feedback component of AMSP provides insights into the use of restricted antimicrobials. This component should be considered by hospitals for inclusion in their program on an ongoing basis even if limited for a few drugs and in few areas of the hospital.

## Introduction

Antimicrobial resistance (AMR) has emerged as a major public health problem all over the world with India as one of the worst affected nations. It is predicted that AMR will lead to 10 million deaths annually by 2050 (1). Excessive use of antimicrobials in humans and animals is the main driving force behind AMR (2). This necessitates the rational use of antimicrobials in the community and effective antimicrobial stewardship programs in hospitals are needed to solve this problem of AMR. Our centre also experiences high rates of resistance in nosocomial gram-negative pathogens with carbapenem resistance rates of 50% and emerging colistin resistance (3). Hence, a need for an urgent up-gradation of the existing Antimicrobial Stewardship Program (AMSP) was felt. We report here our experience with the design, implementation and results of the prospective audit and feedback based component of the AMSP at our hospital.

## Material & Methods

This study reports AMSP data from a private tertiary care hospital with 750 beds including 200 intensive care beds commissioned in 2009. It is accredited by the National Accreditation Board of Hospitals, Joint Commission International and College of American Pathologists. The hospital has an antimicrobial stewardship committee (AMSC) consisting of a clinical microbiologist, infectious disease specialist, clinical pharmacist and infection control nurse. The pre-existing components of the hospital AMSP included generation of antibiogram, formulation/ education and dissemination of antibiotic policies for surgical prophylaxis, community and hospital acquired infections and auditing antibiotics for surgical prophylaxis. We added audit and feedback component about use of restricted antimicrobials since Jan 2018 which is ongoing. These restricted antimicrobials included colistin, polymyxin B, tigecycline, intravenous (IV) minocycline, IV fosfomycin, daptomycin & echinocandins (caspofungin, micafungin & anidulafungin). Use of ceftazidime avibactam was audited from September 2019 onwards. These antimicrobials were chosen as they are the last leg of defence against extremely drug resistant gram negative, gram positive pathogens and Candida and there is as a dire need to preserve their efficacy. The study was approved by the institutional research and ethics committee of Kokilaben Dhirubhai Ambani Hospital and Medical Research Institute (IEC Code: C-3/11/2019).

The prescription of any of these antimicrobials necessitated the filling of an antimicrobial justification form (Supplemental file 1) which was then sent to the antimicrobial stewardship committee. These forms were tallied with a daily indent list from the pharmacy of restricted antimicrobials and any missing forms were requested to be submitted. At 48-72 hours from the time of prescription, the AMSC reviewed the appropriateness of the restricted antimicrobials on the basis of the index patient’s clinical history and course, previous antimicrobial history and results of investigations and cultures. The committee opined whether the use of the antimicrobial is justified/unjustified and about how therapy can be optimized for a particular infection. The same was communicated to the treating clinician and the patients were followed up to assess the compliance to the recommendation of the stewardship committee. Those prescriptions where the restricted antimicrobials were stopped and where the patient died or left the hospital against medical advice earlier than 48 hours were marked as not applicable for review.

The data was collated and analysed monthly to include the number of antimicrobials prescribed, no of patients receiving restricted antimicrobials in absolute numbers and as percentage of the number of monthly admissions, percent prescriptions not applicable for review, percent use unjustified and percentage compliance to the stewardship committee recommendations. Days of therapy (DOT) of the restricted antimicrobial was calculated per 1000 patient days. Rates of colistin resistance in central line associated blood stream infections for years 2018-2019 was calculated and compared with previous years (3). These data were also communicated during the hospital infection control meetings.

Normality of data was assessed using Shapiro Wilk test. For normally distributed data, difference between 2018 and 2019 was analysed using Independent sample T test and data was presented as Mean±SD.

## Results

A total of 2397 restricted antimicrobials in 1366 patients were prescribed in the study period. While in year 2018, 9 forms were missing, 100% of forms were obtained in 2019. Twenty five percent of prescriptions were not applicable for review; further analysis is limited to 1801 prescriptions for 1045 patients (Table 1). Sixty percent were from the adult ICU with rest divided almost equally between the Paediatric ICU, Bone marrow transplant ICU and the Wards with no significant difference between years 2018-2019 (Figure 1).

**Table 1:** Study results from 2018 and 2019.

|  | **Year 2018** | **Year 2019** | **Total** |
| --- | --- | --- | --- |
| Newly admitted patients | 36549 | 39564 | 76113 |
| Number of restricted antimicrobials prescribed | 1203 | 1194 | 2397 |
| Number of forms received | 1194 | 1194 | 2388 |
| % compliance | 99.3% | 100% | 99.6% |
| Number (%) of forms NA for review | 305 (25.5%) | 282 (23.6%) | 587 (24.6%) |
| Number of forms analysed (%) | 889 (74.5%) | 912 (76.4%) | 1801 (75.4%) |
| Number of patients who were on restricted antimicrobials (of forms analysed) | 512 | 533 | 1045 |
| % of newly admitted patients initiated on restricted antimicrobials | 1.4% | 1.3% | 1.37% |
| Total days of therapy/1000 patient days | 41 | 42 | 41.5 |
| Number (%) use Unjustified | 108 (12.1%) | 123 (13.5%) | 231 (12.5%) |
| Number (%) compliance to recommendations | 93 (86.1%) | 113 (91.9%) | 206 (89.2%) |

**Figure 1:**
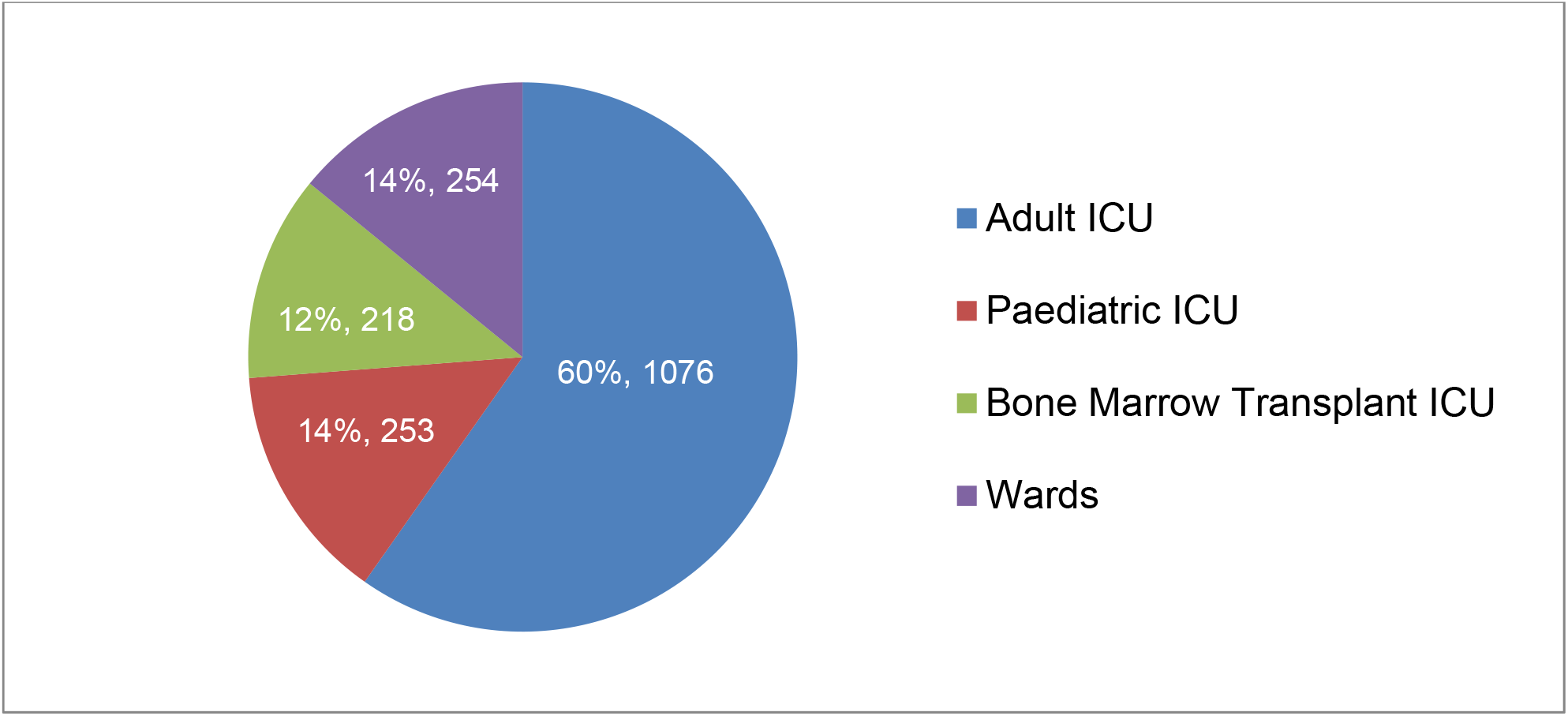
Place where restricted antimicrobials were used (2018 and 2019)

Overall, around 1.4% of admitted patients were put on restricted antimicrobials. The total days of therapy (DOT) were 41.5/1000 inpatient days. Unjustified use of antimicrobials was reported in 13% and recommendation of the AMSC for de-escalation were accepted in 89% by the treatment team. There was no significant difference between the years 2018-2019 in any of the study outcomes (Table 1). Similarly there was no significant difference between antimicrobial days of therapy (DOT) of the restricted antimicrobials between 2018 and 2019 (Table 2). The colistin susceptibility rates remained stable compared to the previous years (Table 3).

**Table 2:** Antimicrobial days per 1000 inpatient days for Echinocandins, Polymyxins (Colistin and Polymyxin), Tetracyclines and Fosfomycin.

|  | **2018** | **2019** | **P value** |
| --- | --- | --- | --- |
| **Echinocandins** | 5.1±1.9 | 5.7±2.1 | 0.566 |
| **Polymyxins** | 21±5.9 | 22±6.8 | 0.729 |
| **Tetracyclines** | 11.4±2.3 | 10.7±3.5 | 0.607 |
| **Fosfomycin** | 3.3±1.6 | 2.6±1.6 | 0.360 |

Data presented as Mean±SD

**Table 3:** Colistin susceptibility (%) of isolates from CLABSI (2011-2019)

|  | **2011** | **2012** | **2013** | **2014** | **2015** | **2016** | **2017** | **2018** | **2019** |
| --- | --- | --- | --- | --- | --- | --- | --- | --- | --- |
| ***Enterobacteriaceae*** | 100 | 98 | 91 | 96 | 96 | 85 | 78 | 100 | 93 |
| ***Acinetobacter* and**<br>***Pseudomonas*** | 100 | 100 | 94 | 100 | 97 | 100 | 94 | 100 | 98 |

## Discussion

We conclude that a prospective audit and feedback based AMSP provides a good insight into the restricted antimicrobial use patterns across the hospital. We were able to get near 100% compliance to receipt of the justification forms. In our observations, the levels of unjustified use (∼13%) was lower, while the near 90% compliance to recommendations of the AMSC is superior to other published studies (4,5). While our data did not show any decline in the use of antimicrobials in the 2nd year, it didn’t rise either. It was also heartening to see that the colistin resistance rates did not rise as compared to previous years. The program also helped in establishing a channel of communication with physicians and helped in optimizing therapy for multi-drug resistant organisms in many instances, an outcome we did not measure.

Prospective audits in ASMP albeit labor-intensive are known to have greater acceptance amongst clinicians as compared to formulary restriction (6). An evaluation of AMSP in India (2008 – 2019) was done by Arzoo et al. (7); the authors reported that the use of prospective audit as an AMSP component was seen in only two studies. Rupali et al conducted prospective audit and feedback study similar to ours in 2 intensive care units of a tertiary-care hospital. The authors reported a significant reduction in antimicrobial use from 831.5 during baseline to 717 DOT in the intervention phase, an effect which was sustained in the follow-up phase (713.6 DOT). The authors also reported inappropriate use of antimicrobials in 73.3% prescriptions and acceptance of recommendations in 60.7% of cases. The second study (Ravi et al.), was a prescription audit and survey-based study. The authors reviewed 121 prescriptions at a tertiary care centre located in Kolkata in 2017. The use of anti-infective medications in the prescription audit rose from 62% in 2014 to 69.1% in 2017. The prescription audits of 2014 and 2017 did not demonstrate major difference in the appropriateness of prescriptions on the parameters for choice, dose, duration, route and combination; although numerical decrease in all the parameters were noted.

A decade long ASMP providing assessments to over 7700 antimicrobial prescriptions for patients admitted to the ICU in Toronto (Canada), gave therapy altering suggestions in 36% prescriptions and noted an acceptance rate of 67% (8). The authors stated that the acceptance rate remained stable during the period and have also stated factors associated with higher and lower likelihood of accepting ASMP recommendations. We also report an insignificant but stable improvement in acceptance to ASMP recommendations over the 2-year period.

The strengths of our set up that aid the AMSP include a full-time specialist system, commitment by the top management and involvement of the infection control nurses. The greatest challenge was that the program was entirely manual and person driven/ dependent. An antimicrobial stewardship software which can be seamlessly integrated with the hospital information system (HIS) would greatly assist our’s and any AMSP.

At certain times non availability of members of the AMSC led to delayed feedback about the inappropriateness of the prescription leading to excess use of inappropriate antimicrobials. While the decision about the justification was black and white in most cases, in some it was difficult to compartmentalize between justified and unjustified.

The single centre nature of the study limits the generalizability. The second limitation is the list of antimicrobials assessed by us were limited and did not include commonly used drugs like carbapenems, vancomycin & teicoplanin. This limitation can possibly explain the lower percentage of antimicrobial use and unjustified use in our study. Therefore, auditing all antimicrobial use in the hospital and its appropriateness is best done as a point prevalence study (9). Third we did not audit the use of multiple antimicrobials, duration of therapy, IV to oral switch, drug doses and antibiotic use in emergency area. We aim to expand the scope of the program by auditing the use of more antimicrobials at least in some areas of the hospital and duration of therapy with antimicrobials.

We recommend that all hospitals should add a prospective audit and feedback component to their AMSP program even if it is for a few drugs and in few areas of the hospital.

## Data Availability

Data is archived with the infection prevention and control department of Kokilaben Hospital

https://www.kokilabenhospital.com

## Acknowledgement for financial support & specific scientific contribution only

The study has not received any financial support from any external funding agencies. We acknowledge all physicians and nurses from our hospital for their support. We also acknowledge the Hospital Information System team and hospital pharmacy for assistance in extracting data on antimicrobial consumption.

